# An updated molecular diagnostic for surveillance of *tetM* in *Neisseria gonorrhoeae*

**DOI:** 10.1101/2024.08.26.24312240

**Authors:** Samantha G. Palace, Jordan A. Reyes, Eric Neubauer Vickers, Aishani V. Aatresh, Wei Shen, Zamin Iqbal, Yonatan H. Grad

## Abstract

Doxycycline post-exposure prophylaxis (doxy-PEP) for sexually transmitted bacterial infections reduces the risk of syphilis and chlamydia, but effectiveness against gonorrhea is variable, likely attributable to varying resistance rates. As doxy-PEP is incorporated into clinical practice, an urgent unanswered question is whether increased doxycycline use will drive tetracycline-class resistance in *Neisseria gonorrhoeae*. Here, we report an updated RT-PCR molecular diagnostic to detect the *tetM* gene that confers high-level tetracycline resistance in *N. gonorrhoeae*.

## Text

Doxycycline post-exposure prophylaxis (doxy-PEP) for sexually transmitted bacterial infections reduces the risk of syphilis and chlamydia, while effectiveness against gonorrhea is variable (1-3), likely attributable to varying resistance rates. High-level resistance to tetracycline in *Neisseria gonorrhoeae*, the agent of gonorrhea, is estimated at 10% prevalence in the United States (4) and up to 70-100% in some populations (5-7). As doxy-PEP is incorporated into clinical practice, an urgent unanswered question is whether increased doxycycline use will drive tetracycline-class resistance in *N. gonorrhoeae*. However, current culture-based surveillance of *N. gonorrhoeae* antimicrobial susceptibility may not be sufficient to quickly detect changes in the prevalence of tetracycline resistance because surveillance programs collect and test a modest number of isolates (e.g. (8)). By contrast, molecular diagnostics for tetracycline resistance would enable the use of remnants from nucleic acid amplification tests (NAATs), the most widely used diagnostic, and can confidently detect changes in population-level resistance more quickly. An accurate molecular diagnostic for high-level tetracycline resistance thus has public health value (9).

High-level tetracycline resistance in *N. gonorrhoeae* is conferred by TetM, a plasmid-encoded ribosomal protection protein. Although primers for PCR-based detection of the *tetM* gene have been reported (10, 11), the existing reverse primer, originally designed from the *Ureaplasma urealyticum tetM* sequence, has poor predicted sensitivity for *N. gonorrhoeae tetM* sequences: only 992 of 2223 *N. gonorrhoeae tetM* sequences deposited in PubMLST (Supplemental File S1) (12) contain an exact match for this primer. We designed a new reverse primer (Supplemental Materials, Table S1) that recognizes all 2223 *N. gonorrhoeae tetM* sequences tested, and that, when used in combination with the reported forward primer from Ison *et al*., is predicted to successfully amplify 2221 of 2223 *N. gonorrhoeae tetM* sequences (or all 2223 sequences if tolerating a single nucleotide mismatch).

To test this primer set, we assembled a panel of 13 diverse clinical isolates comprising 6 *tetM*+ and 7 *tetM*-strains (Supplemental Materials, Figure S1, Table S2). Genomic DNA was extracted, and *tetM* primers were tested for specific and sensitive amplification against this panel (Supplemental Materials, Methods), using *porA* as a reference amplicon (13). As expected, *tetM* amplification segregates clearly with the presence or absence of *tetM* in all strains (Figure 1A). Accurate discrimination was maintained over a range of template concentrations (Figure 1B) to a limit of approximately 28 copies of the gonococcal genome per reaction, at which point the reaction approaches its sensitivity limit (Supplemental Materials, Figure S2).

**Figure 1.**
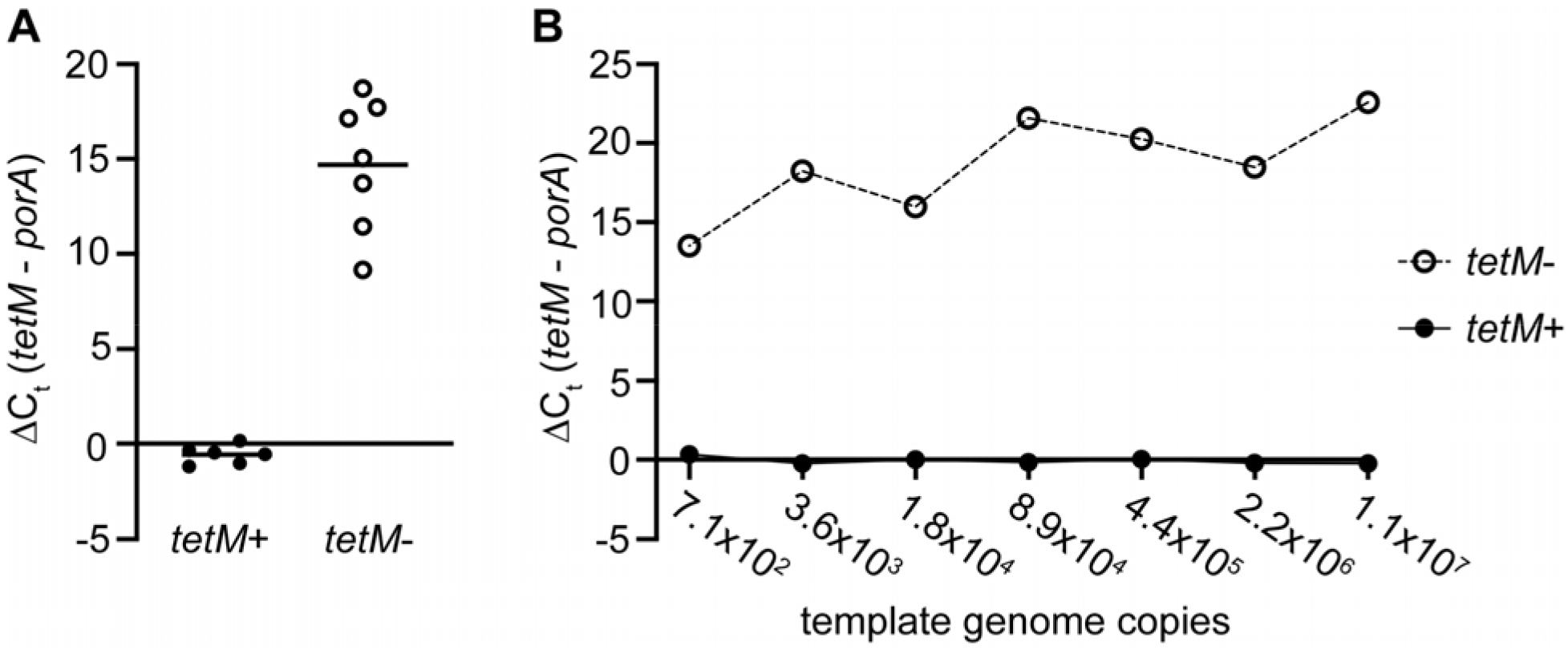
RT-PCR-based detection of *tetM* in a diverse strain panel. Mean cycle threshold (C_t_) of *tetM* amplification, baseline corrected by *porA* amplification, for **(A)** 2.5 ng genomic DNA purified from strains with *tetM* (n=6, left) and without *tetM* (n=7, right); and **(B)** over a variety of template concentrations for one representative strain each with *tetM* and without *tetM*. Amplification was monitored via SYBRGreen fluorescence on a QuantStudio™ 7 Flex Real-Time PCR system (see Supplemental Methods). Shown: mean ΔC_t_ from three technical replicates per reaction; data representative of at least two independent experiments.

The sensitivity and specificity of this test for clinical samples, and importantly for GC-positive NAAT remnants, requires further characterization. Because *tetM* in *N. gonorrhoeae* originated from a streptococcal transposon (14), highly similar *tetM* sequences are found in other organisms, creating a risk of false positive *tetM* detection. Comparing *tetM* positivity rates in GC-positive and GC-negative NAAT specimens will be an important step in characterizing the expected false positive rate. The use of *porA* primers for normalization may also mitigate this issue. Although the data here were generated using dye-based (SYBR Green) RT-PCR detection, probe-based detection may also improve specificity.

## Data Availability

All data produced in the present work are contained in the manuscript

## Ethics statement

Clinical isolates were obtained from the CDC and FDA Antimicrobial Resistance Isolate Bank, NCIP panel (15) and from previously published studies (16, 17). The original collection of these isolates was deemed not human subjects research by the Center for Disease Control and Prevention’s Office of the Associate Director for Science for the Gonococcal Isolate Surveillance Program (GISP/eGISP) (16, 18) or by the institutional review boards at the University of California San Diego, University of Washington, and Harvard T.H. Chan School of Public Health (17).

