## Supplemental materials for "An updated molecular diagnostic for surveillance of *tetM* in *Neisseria gonorrhoeae*"

**Supplemental information**

**Methods**

**Bacterial strains and genomic DNA isolation.** A panel of 13 sequenced *Neisseria gonorrhoeae* strains with (n=6) and without (n=7) *tetM* was selected to span the phylogenetic diversity of our strain collection (Figure S1, Table S2). Strains were cultured on GCB agar (Difco) with Kellogg’s supplements (1) at 37°C with 5% CO­_2_. Overnight growth was collected with a sterile swab and genomic DNA was isolated with a PureLink™ Genomic DNA Mini Kit (Invitrogen) and eluted in nuclease-free water. Yield was quantified with a Qubit 3.0 fluorometer (Invitrogen) and DNA was diluted in nuclease-free water for RT-PCR.

**RT-PCR.** Primers used in this study are presented in Table S1. Purified genomic DNA was added at the specified concentrations to PowerUp SYBRGreen Master Mix (Appied Biosystems) with 0.5μM primers porA_F and porA_R (for *porA* amplification) or tetM_F4 and tetM_R5 (for *tetM* amplification). 6.25 μL reactions were run in MicroAmp™ Optical 384-well reaction plates (Applied Biosystems) on a QuantStudio™ 7 Flex Real-Time PCR system (Applied Biosystems) on fast cycling settings for 20s at 95°C, followed by 40 cycles of 1s at 95°C and 20s at 63°C. Melt curves were performed to assess product purity. Cycle thresholds were calculated by QuantStudio™ 7 Flex software. Baseline correction of *tetM* amplification by *porA* amplification was performed by reporting delta(C_t_) = *tetM*(C_t_) – *porA*(C_t_).

**Supplemental file S1.** Alignment of *tetM* sequences deposited in PubMLST for *Neisseria gonorrhoeae*.

**Table S1.** Primers in this study.

| Primer | Sequence | Source |
| --- | --- | --- |
| porA_F | CCGGAACTGGTTTCATCTGATT | (2) |
| porA_R | GTTTCAGCGGCAGCATTCA | (2) |
| tetM_F4 | GGCGTACAAGCACAAACTCG | (3) |
| tetM_R4 | TCTCTGTTCAGGTTTACTCG | (3) |
| tetM_R5 | GTTCAGATTCGGTAAAGTTCGTC | This study |

**Table S2.** Characteristics of *N. gonorrhoeae* strains in this study. MLST, NG-MAST, NG-STAR, and NG-STAR clonal complexes were determined with pyngoST (4).

| Strain | *tetM* | MLST | NG-MAST | NG-STAR | NG-STAR_CC | Accession | Reference |
| --- | --- | --- | --- | --- | --- | --- | --- |
| CCC011 | N | 13736 | - | 832 | CC3035 | SRR16683507 | (5) |
| CCC015 | Y | 1587 | 12818 | 719 | CC427 | SRR16683503 | (5) |
| GCGS0363 | Y | 10932 | 19091 | 399 | CC390 | ERR855137 | (6) |
| GCGS0440 | N | 1601 | - (untyped POR) | - (untyped *penA*) | - | ERR1067717 | (6) |
| GCGS0605 | N | 1579 | 5 | 139 | CC442 | ERR1067711 | (6) |
| GCGS0620 | Y | 10932 | 4321 | 383 | CC390 | ERR1067811 | (6) |
| GCGS0870 | N | 10316 | - (untyped POR) | 685 | CC3165 | ERR1067778 | (6) |
| JJJ012 | Y | 7371 | 10495 | - (untyped *porB*) | - | SRR16683802 | (5) |
| NCIP-05 | N | 6962 | 1063 | 391 | CC3087 | SRR8833292 | (7) |
| NCIP-06 | N | 8149 | 1319 | 688 | CC84 | SRR8833294 | (7) |
| NCIP-08 | N | 1893 | - (untyped POR) | - (untyped *mtrR*) | - | SRR8833288 | (7) |
| NCIP-09 | Y | 1600 | 2194 | 2784 | CC514 | SRR8833289 | (7) |
| NCIP-11 | Y | 8152 | - | - | - | SRR8833290 | (7) |

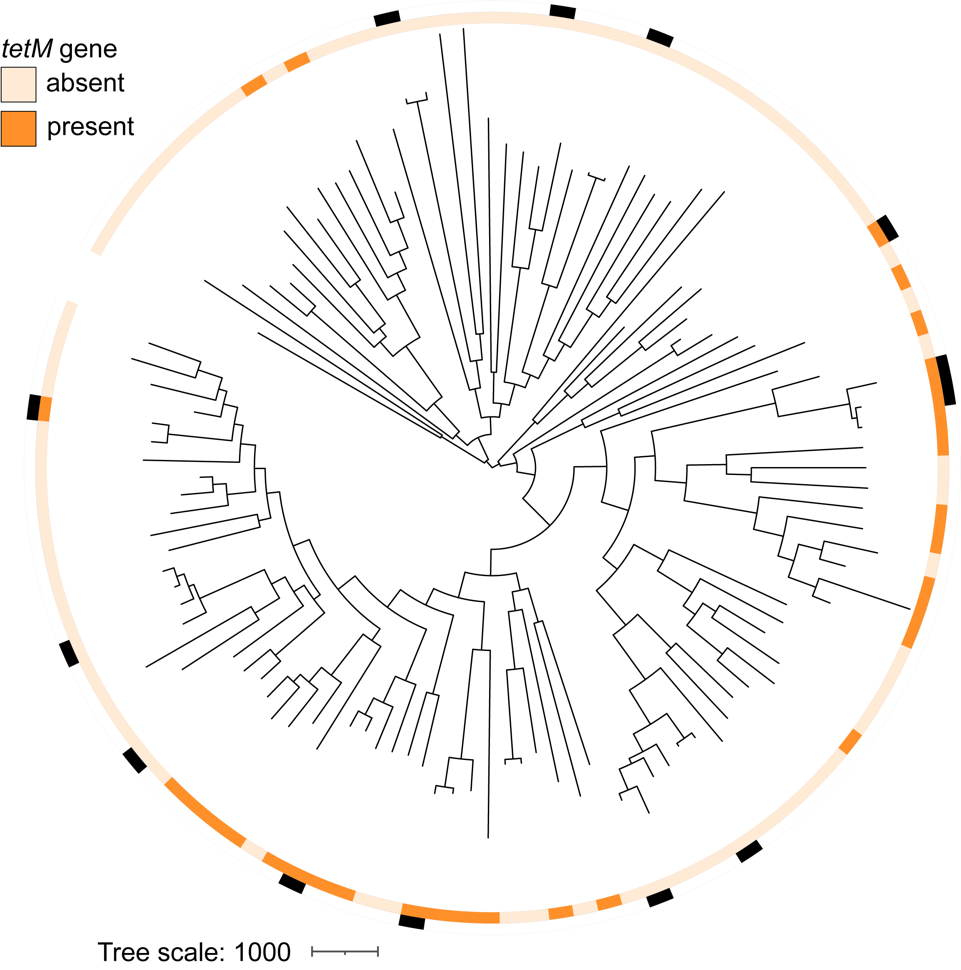

**Figure S1.** Phylogenetic distribution of the *tetM* strain panel. Strains used in this study (marked in black, outer ring) were placed on a phylogeny of 100 isolates that represent the maximal genetic diversity of our laboratory’s collection of *N. gonorrhoeae* isolates. Inner ring: presence (dark orange) or absence (tan) of the *tetM* gene. Phylogeny was created from pseudogenomes with recombinant regions masked by Gubbins (8) using RAxML-NG (9) with the GTRGAMMA nucleotide substitution model and visualized using iTOL (10).

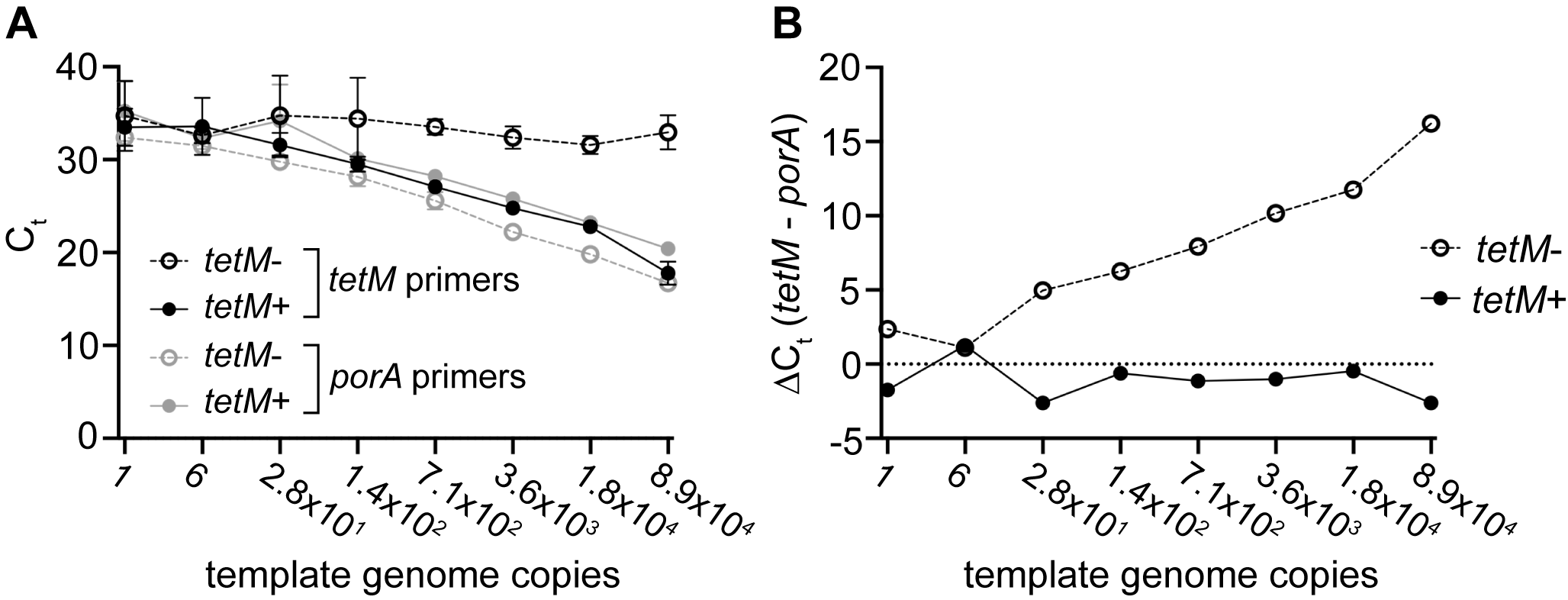

**Figure S2.** Limit of detection of the *tetM* primers. RT-PCR was run with five-fold dilutions of genomic DNA from CCC011 (*tetM*-) or CCC015 (*tetM*+) to determine the limit of detection. **(A)** As the template amount decreases towards the assay’s limit of detection, reduced amplification (higher C­­­_t_ values) for both primer pairs results in **(B)** a reduced difference between the *porA*-corrected *tetM* amplification signal for the *tetM*+ and *tetM*- strains. Shown: C_t_ mean and standard deviation (A) or mean ΔC_t_ (B) from four technical replicates per reaction; data representative of at least two independent experiments.
